# Connecting diet and disease: Using Mendelian randomisation to bridge the gap

**DOI:** 10.64898/2026.09.02.26362020

**Authors:** Benedita Deslandes, Laura J Corbin, Lucy J Goudswaard, Meda R Sandu, Matthew A Lee, Rhona A Beynon, Lucy McGeagh, George Davey Smith, Naveed Sattar, Michael EJ Lean, Roy Taylor, J Athene Lane, Nicholas J Timpson, Richard M Martin, George Richenberg, Marc J Gunter, James Yarmolinsky, Françoise Koumanov, Javier T Gonzalez, Rebecca C Richmond, Emma E Vincent

**Affiliations:** MRC Integrative Epidemiology Unit at the University of Bristol, Bristol, United Kingdom; Population Health Sciences, Bristol Medical School, University of Bristol, Bristol, United Kingdom; The National Institute for Health Research (NIHR) Bristol Biomedical Research Centre, University Hospitals Bristol and Weston NHS Foundation Trust and University of Bristol, Bristol, United Kingdom; International Agency for Research on Cancer, WHO, Lyon, France; Oxford Institute of Applied Health Research (OxInAHR), Oxford Brookes University, Oxford, UK; Institute of Health and Wellbeing, and Institute of Cardiovascular and Medical Science, University of Glasgow, Glasgow, UK; Human Nutrition, School of Medicine, Dentistry and Nursing, College of Medical, Veterinary and Life Sciences, University of Glasgow, Glasgow, UK; Newcastle Magnetic Resonance Centre, Translational and Clinical Research Institute, Campus for Ageing and Vitality, Newcastle University, Newcastle upon Tyne, UK; Bristol Trials Centre, Bristol Medical School, University of Bristol, Bristol, United Kingdom; Cancer Epidemiology and Prevention Research Unit, School of Public Health, Imperial College London, UK; Department of Epidemiology and Biostatistics, School of Public Health, Imperial College London, UK; Department for Health, Centre for Nutrition Exercise and Metabolism, University of Bath, Bath, UK; Translational Health Sciences, Bristol Medical School, University of Bristol, Bristol, UK

**Author notes:** Joint senior authors (Rebecca C Richmond, Emma E Vincent).

## Abstract

Establishing causality in nutrition research is challenging. While randomised controlled trials (RCTs) provide robust evidence, long-term dietary intervention studies with disease endpoints are often impractical. Short-term RCTs can instead identify intermediate traits that may lie on the causal pathway between diet and disease. Mendelian randomisation (MR) is an epidemiological approach that uses genetic variants as proxies for modifiable exposures to estimate the effects of lifelong differences in exposure on disease risk. However, the utility of MR is limited for complex dietary patterns because genetic variants typically reflect biological mechanisms rather than specific diets.

We propose a two-step framework integrating dietary RCTs with MR to infer potential lifetime effects of dietary interventions. First, RCT data identify molecular traits altered by an intervention. Second, MR evaluates whether these traits are associated with long-term disease risk.

We demonstrate this framework using the Diabetes Remission Clinical Trial (DiRECT), which measured circulating proteins and diabetes remission. Using protein data alone, 216 of 4,601 proteins changed following the intervention (step 1), and 10 were associated with diabetes risk using MR (step 2). We then compared these MR estimates with observed protein-remission associations from DiRECT. The broad agreement between the two (r≈−0.645, R^2^=0.416) supports this framework as a useful approach for estimating long-term effects of dietary interventions.

## Introduction

Nutrition science faces unique challenges in establishing causality due to the complex, time-varying, and intercorrelated nature of dietary exposures (1). Strongest causal inference is obtained through the combination of randomisation, exogenous manipulation and replication, all of which can be incorporated within randomised controlled trials (RCTs). However, conducting long-term dietary intervention RCTs specific to disease development is often impractical due to timescale, cost, the inherent complexity of nutrition research, as well as the ethical challenges associated with maintaining participants on potentially harmful diets for prolonged periods (2). Therefore, many dietary RCTs are often constrained to shorter durations and focus on short-term, or intermediate, outcomes. Results from these studies may not adequately capture the relationship between dietary exposures and long-term disease outcomes, particularly where dietary changes need to be sustained over longer periods to confer an effect.

Mendelian randomisation (MR) is a study-design which, under certain assumptions, can strengthen causal inference between traits (4). MR uses genetic variants, typically single nucleotide polymorphisms (SNPs), as instrumental variables for modifiable exposures to estimate causal effects on outcomes. Given that genetic variants are fixed at conception and randomly assorted, MR is generally less susceptible to confounding and reverse causation than conventional observational studies (**Figure 1**) (3,4). One advantage of MR is its ability to provide insight into the effects of lifelong differences in exposure, which may not be captured in shorter-term intervention studies. It is often inappropriate to perform MR studies of nutritional exposures because robust genetic proxies are difficult to identify, and genetic predisposition to a dietary behaviour may not recapitulate the biological effects of adopting that behaviour through environmental or behavioural change (1,5). In other words, MR assumes that changing an exposure through genetics has similar biological effects to changing it through an intervention, such as a dietary modification. This is known as the gene-environment equivalence assumption (6).

**Figure 1.**
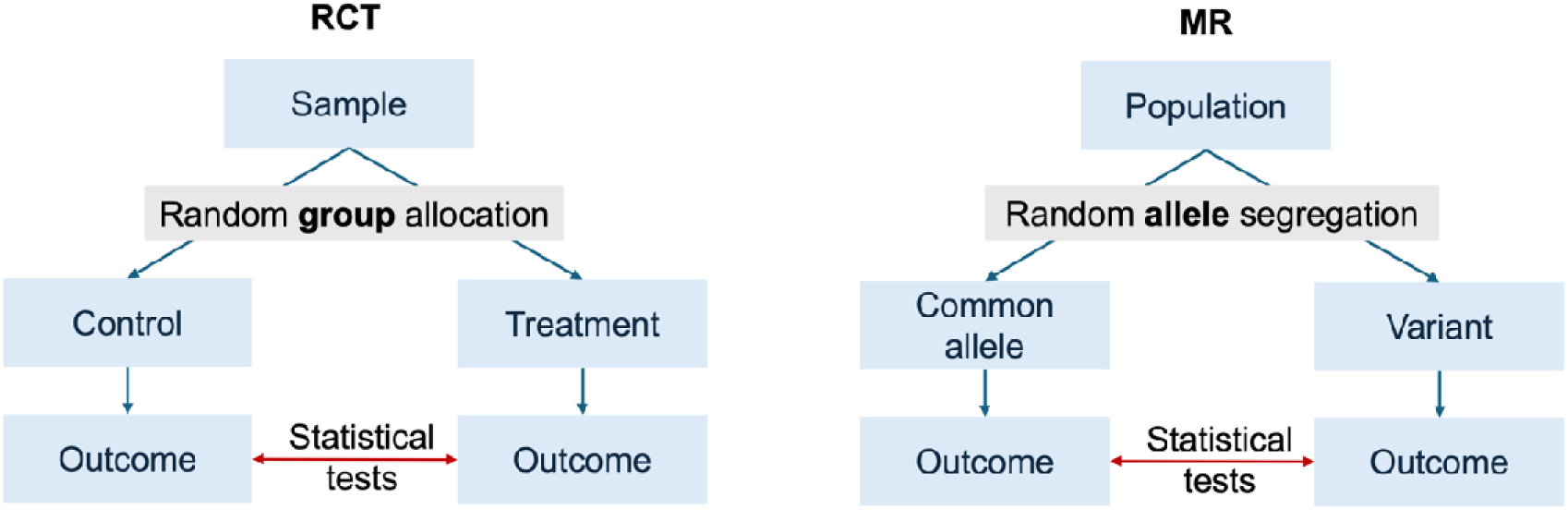
Conceptual comparison of randomised controlled trials (RCT) and Mendelian randomisation.

Consequently, both dietary intervention RCTs and MR have important but distinct limitations when used independently to infer causal relationships between diet and long-term health outcomes. Here, we propose a methodological framework that addresses the complementary limitations from both study designs by focusing on intermediate molecular traits that change in response to the dietary interventions, and importantly which may lie on the causal pathway to disease. To demonstrate the utility of this two-step approach, we use data from the Diabetes Remission Clinical Trial (DiRECT) (7). DiRECT evaluated the effect of a structured dietary weight-loss programme (n=149), compared to best-care practice by dietary guidelines (n=149), on remission of type 2 diabetes (T2D) at 1-year follow-up. DiRECT provides an ideal opportunity to test our proposed framework because it measured both intervention-associated molecular changes and a clinically relevant outcome (T2D remission), enabling framework-derived predictions to be validated against observed remission outcomes.

## Methodological approaches

### Randomised controlled trials

In an RCT, participants are randomly allocated to one of several study arms to receive an intervention (*e.g.* dietary modification) or a control/comparator condition. The most commonly used analytical approach is intention-to-treat (ITT) analysis. ITT analyses compare participants according to their random allocation, regardless of adherence, therefore preserving the benefits of randomisation (8). In this respect, ITT analyses are conceptually analogous to MR, where individuals are effectively randomised to different levels of an exposure based on their genetic variants, irrespective of subsequent behavioural or environmental modification. Key challenges faced with dietary modification RCTs are summarised in **Table 1**.

**Table 1.**
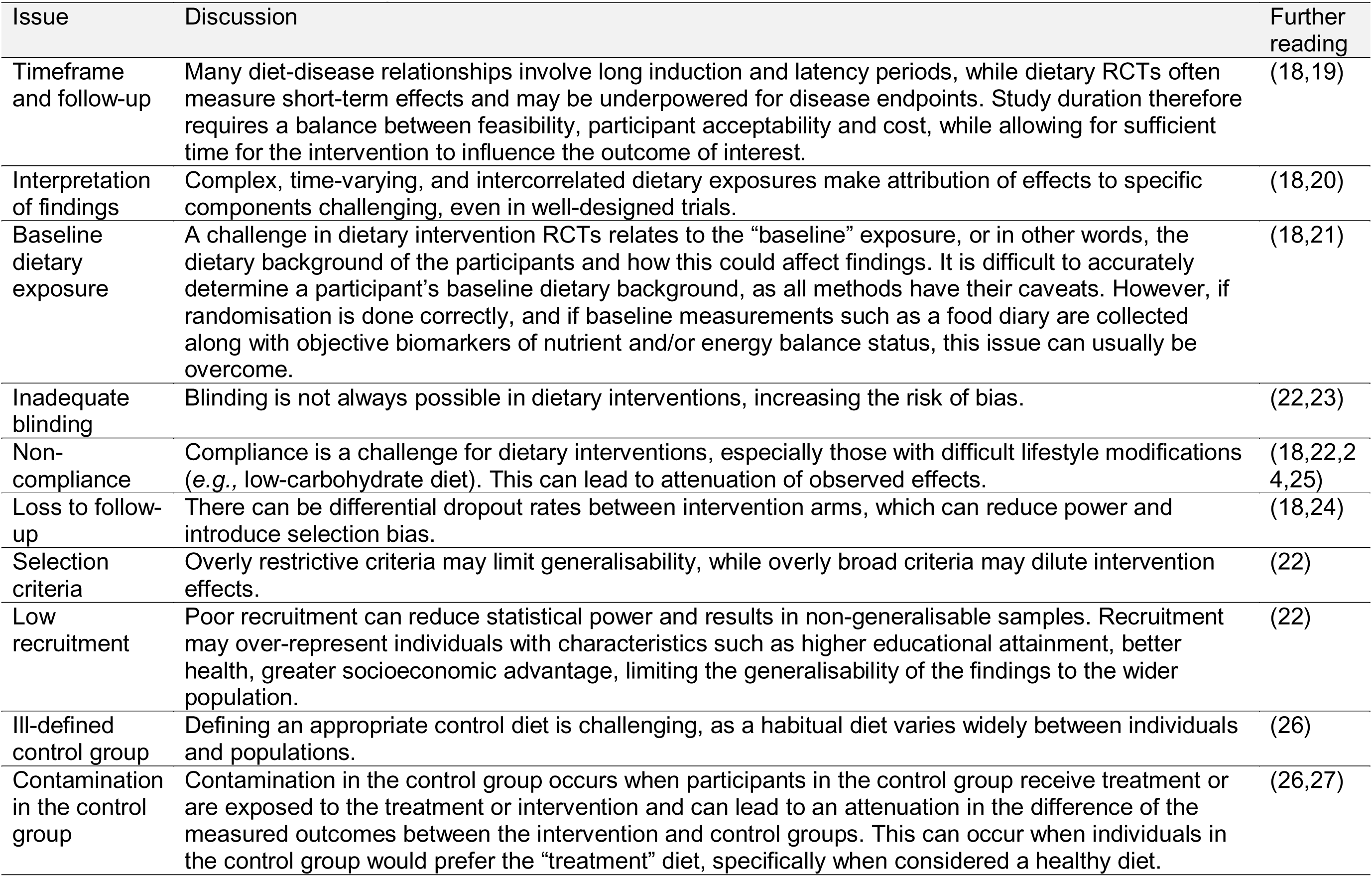
Common issues in dietary intervention randomised controlled trials.

Often, surrogate endpoints are used in feasibility studies or trials with relatively short follow-up (9). These endpoints can include molecular features such as circulating proteins, which may respond more rapidly than clinical outcomes and provide insight into potential biological mechanisms (10). However, while these markers may lie on the pathway between an intervention and disease, they do not necessarily reflect causal effects on clinical outcomes. As a result, additional approaches are required to determine whether these molecular changes are likely to be causally relevant. MR offers one such approach to assess the causal relevance of these intermediate traits.

### Mendelian randomisation

MR uses genetic variants as proxies for modifiable exposures to investigate potential causal relationships between exposures and outcomes (**Figure 1**). Although MR is often described as a natural experiment, the analogy with randomisation is most directly applicable to within-family designs, where parental alleles are randomly transmitted to offspring. In population-based MR, genetic instruments may be associated with familial, social or environmental factors, which may be particularly relevant to dietary behaviours (3). Nevertheless, genetically influenced differences in an exposure can provide an opportunity to investigate its potential effect on an outcome (3,4). MR relies on three core assumptions (**Figure 2**): (i) the genetic variant(s) are robustly associated with the exposure; (ii) the genetic instrument is independent of confounders of the instrument-outcome association; and (iii) the genetic variant(s) influence the outcome only through their effect on the exposure (3). For example, a genetic variant associated with circulating protein levels should influence disease risk through the protein itself rather than through an alternative biological pathway. Detailed discussions and definitions of these assumptions and their implications have been described extensively elsewhere (11,12).

**Figure 2.**
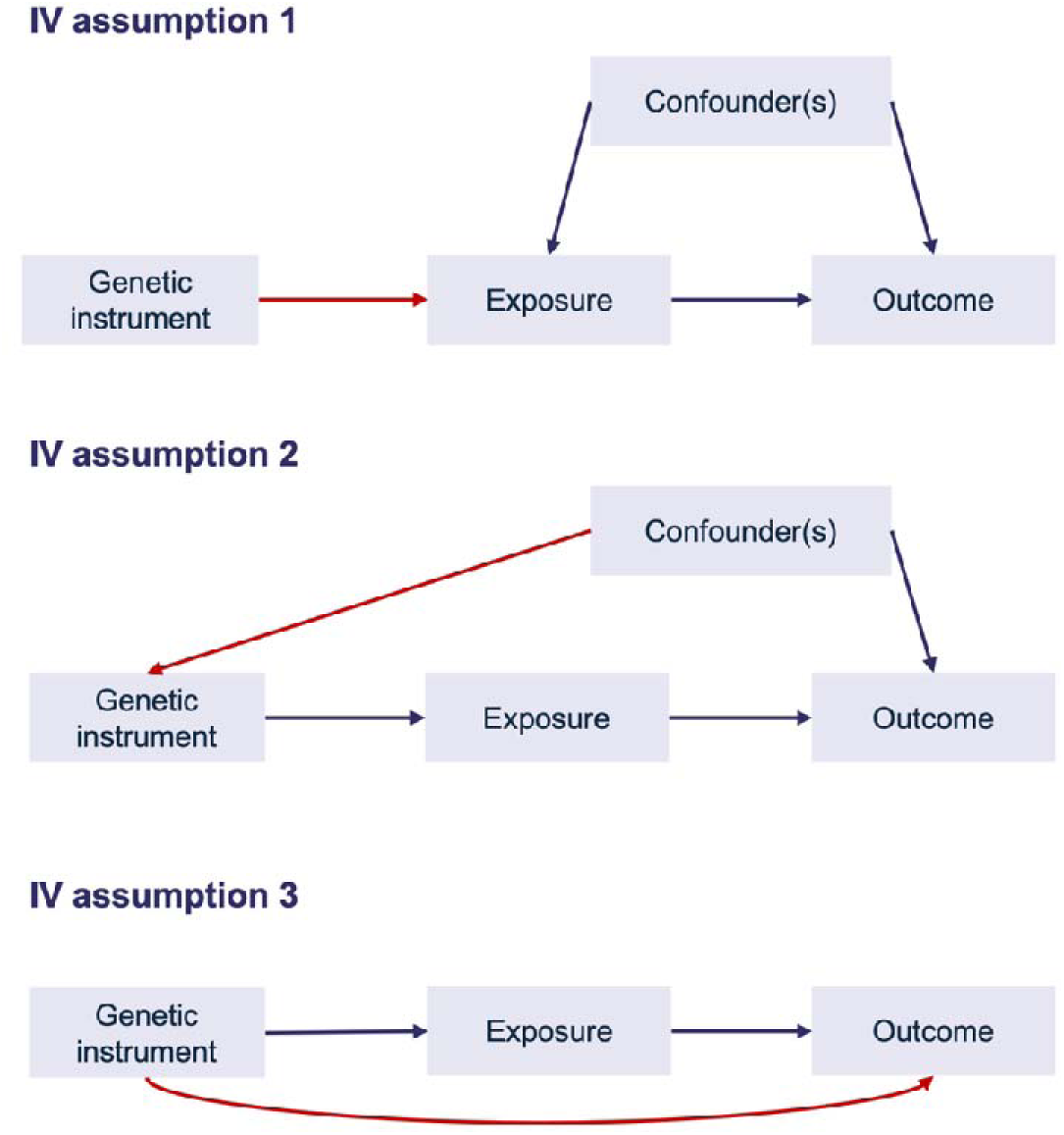
Mendelian randomisation three core assumptions. Assumptions highlighted in red: (i) the genetic variant(s) are robustly associated with the exposure; (ii) the genetic variant(s) are independent of factors that could confound the association between the genetic instrument and outcome; and (iii) the genetic variant(s) influence the outcome only through their effect on the exposure.

MR can be conducted using either individual or summary-level data. Individual-level MR uses data where genetic variants, exposures and outcomes are measured in the same participants. In contrast, summary-level MR uses aggregate genetic associations, typically obtained from large genome-wide associations studies (GWAS). Summary-level approaches are now more commonly used because large GWAS datasets are widely available and generally provide greater statistical power. Readers are referred to these publications for a comprehensive overview of MR and the range of methodological approaches available (11–13).

While MR can, in principle, be applied to a wide range of exposures, its application in nutrition research is often limited (1). Whole dietary behaviours and patterns are inherently complex and highly correlated, making it difficult to identify suitable genetic instruments, that is, variants that explain a meaningful proportion of variance in the exposure, for an entire diet rather than a specific nutrient or biomarker (1). Genetic variants identified in dietary GWASs may reflect broader behavioural socioeconomic, or physiological traits, such as adiposity, educational attainment, or general health, rather than dietary intake itself. This can introduce heritable confounding and complicate interpretation of causal estimates (14,15). Consequently, MR has been most informative in relatively simple scenarios, where the exposure of interest is a single, well-defined nutrient, or nutritional biomarker. An example is the use of MR to recapitulate the outcome of the Selenium and Vitamin E Cancer Prevention Trial (SELECT), which investigated selenium supplementation and prostate cancer risk (16,17). In this case, MR analysis suggested that genetically predicted higher circulating selenium levels were associated with potential adverse outcomes, findings which were consistent with the SELECT trial results (17). This and similar examples are summarised in a recent review of MR studies investigating the role of nutritional exposures in cancer outcomes (14). Such analyses also rely on gene-environment equivalence: genetically influenced differences in the intermediate trait are assumed to have comparable downstream effects to changes induced by the dietary intervention (6). Common methodological challenges in MR, particularly when applied to dietary exposures, are summarised in **Table 2**.

**Table 2.** Common issues in summary-level Mendelian randomisation.

| Issue | Discussion | Methods to mitigate | Further reading |
| --- | --- | --- | --- |
| Methodological misapplication and misinterpretation | The validity of MR findings depends on appropriate instrument selection, analytical methods, assessment of assumptions, and careful interpretation of causal estimates. Inappropriate application or interpretation of MR can lead to biased or misleading conclusions. | Attend courses, understand different methods and when they should be applied, choose genetic instruments appropriately, MR-STROBE, triangulation of evidence. | (6,28–31) |
| Weak instrument bias | When genetic instruments have weak associations with the exposure, this results in biased or imprecise causal estimates. | Use adequately powered GWAS to identify genetic instruments; combine multiple independent exposure associated variants where appropriate. | (32–35) |
| Linkage disequilibrium (LD) | Correlated variants may inflate precision or violate instrumental variable assumptions. | LD clumping<br>Colocalisation analyses | (35,36) |
| Population stratification | Genetic structure correlated with exposure or outcome can bias causal estimates. | Ancestry-restricted analyses<br>Adjustment for principal components of genetic ancestry<br>Within-family analyses | (37–39) |
| Horizontal pleiotropy | Genetic variants influence the outcome through pathways other than the exposure of interest, either directly or indirectly. | MR-Egger, weighted median and mode-based MR to assess robustness to horizontal pleiotropy; methods such as CAUSE to account for correlated pleiotropy. | (40–44) |
| Collider bias | Selection or adjustment for variables influenced by the exposure and outcome can induce collider bias and affect genetic associations in MR. | Avoid inappropriate covariate adjustment or selection in GWAS; assess the potential for collider bias and apply correction methods such as Slope-Hunter where appropriate. | (45–50) |
| Sample overlap | Sample overlap between exposure and outcome GWAS can bias causal estimates in the presence of weak instruments. | Use non-overlapping datasets where possible. Apply jackknife or other overlap-correction methods when overlap cannot be avoided | (3,33–35,51–53) |
| Canalisation | Biological compensation or developmental adaptation to genetic variation may alter the effect of genetic instruments on the exposure. | May attenuate or bias causal estimates; no clear method to fully account for this. | (54) |
| Non-linearity | Exposure-outcome relationships may not be linear. | Although several methods (e.g., instrument-free and doubly-ranked approaches) exist, they require large sample sizes, strong assumptions, and are usually only feasible with one-sample MR using individual-level data. | (55–58) |
| Violations of homogeneity or monotonicity | Violations of these assumptions affect the interpretation and generalisability of the causal effect estimated by MR. | Consider homogeneity and monotonicity when interpreting MR estimates. | (59–62) |

### Two-step framework

Our two-step framework leverages the key strengths of RCTs and MR whilst addressing their corresponding limitations. In the first step, data from a short-term dietary intervention RCT is used to identify intermediate traits, *e.g.,* circulating proteins, metabolites, or gene expression in tissue, that change in response to the intervention (**Figure 3, Step 1)**. In the second step, these intermediates are treated as exposures in MR analyses to assess their potential causal effects on long-term health outcomes (**Figure 3, Step 2)**. This approach builds on the previously described two-step epigenetic MR framework (63), but replaces the first MR step with a short-term RCT to identify intervention-responsive intermediate traits. When the exposure being instrumented is a well-defined molecular trait, MR can provide more robust causal evidence than is typically possible for whole dietary patterns.

**Figure 3.**
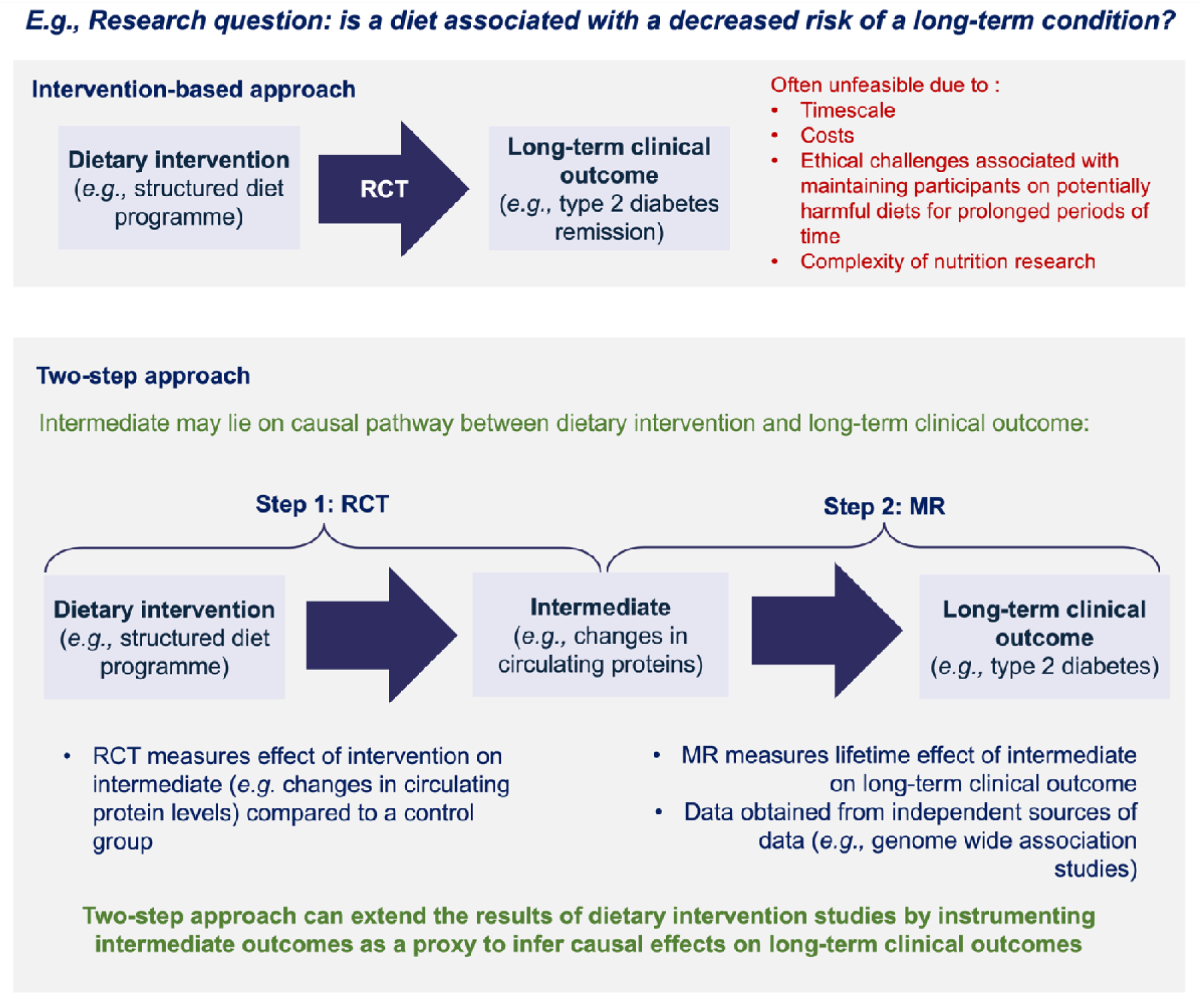
Example of how we can extend the results of small-scale dietary intervention studies by instrumenting intermediate outcomes to infer causal effects on long-term clinical outcomes. MR = Mendelian randomisation; RCT = randomised controlled trial.

To date, we identified three studies that have explicitly applied a similar two-step RCT-MR framework to investigate the relationship between nutrition exposures and disease outcomes, which are summarised in **Table 3**. Both Beynon et al., (64) and Bull et al., (65) utilised randomised controlled designs with comparisons between intervention and control groups, whereas Pietzner et al., (66) examined proteomic changes in response to a short-term fasting intervention without a formal control group, which may limit causal interpretation of the initial step. These studies help us demonstrate the feasibility of the approach and illustrate how short-term molecular responses to dietary interventions can be leveraged to gain insight into long-term disease risk.

**Table 3.** Description of published studies using two-step framework.

| Study | Two-step framework |  |  | Findings |
| --- | --- | --- | --- | --- |
|  | Dietary intervention | Intermediate | Disease outcome |  |
| Beynon et al. (2019) (64) | 6-month intervention with lycopene and green tea in men at risk of prostate cancer | Serum metabolites (pyruvate, valine, acetate, DHA) | Prostate cancer | Lycopene intake altered serum metabolites, particularly reducing pyruvate. MR analysis suggested higher pyruvate levels may increase prostate cancer risk. |
| Bull et al. (2024) (65) | DiRECT trial: structured dietary programme that led to weight loss. | Circulating proteins | Risk of several cancers (colorectal, breast, endometrial, gallbladder, liver and pancreatic) | Intentional weight loss in people recently diagnosed with T2D may modify levels of cancer-related proteins in serum. |
| Pietzner et al. (2024) (66) | 7-day complete caloric restriction in 12 volunteers | Circulating proteins | Metabolic disorders, cardiovascular disease, rheumatoid arthritis | Fasting induces systemic protein-level changes, suggesting potential mechanisms for metabolic and cardiovascular health effects. |

### Practical considerations

Successful application of the two-step framework depends on alignment between the intervention study (Step 1) and the MR analysis (Step 2). When designing a novel RCT, both components should be considered in tandem to ensure that molecular traits measured in the intervention can be evaluated using appropriate genetic instruments and GWAS data. If starting from an existing intervention study, the MR analysis should be designed around the available molecular endpoints, selecting only those traits for which suitable genetic instruments and outcome data exist. Conversely, if designing a novel RCT, it should be designed to measure molecular traits that meet the requirements for downstream causal inference using MR.

### Worked exemplar

We now conduct a worked example using data from the DiRECT trial (7). **Figure 4** illustrates the methodological steps taken to demonstrate the validity of the two-step framework.

**Figure 4.**
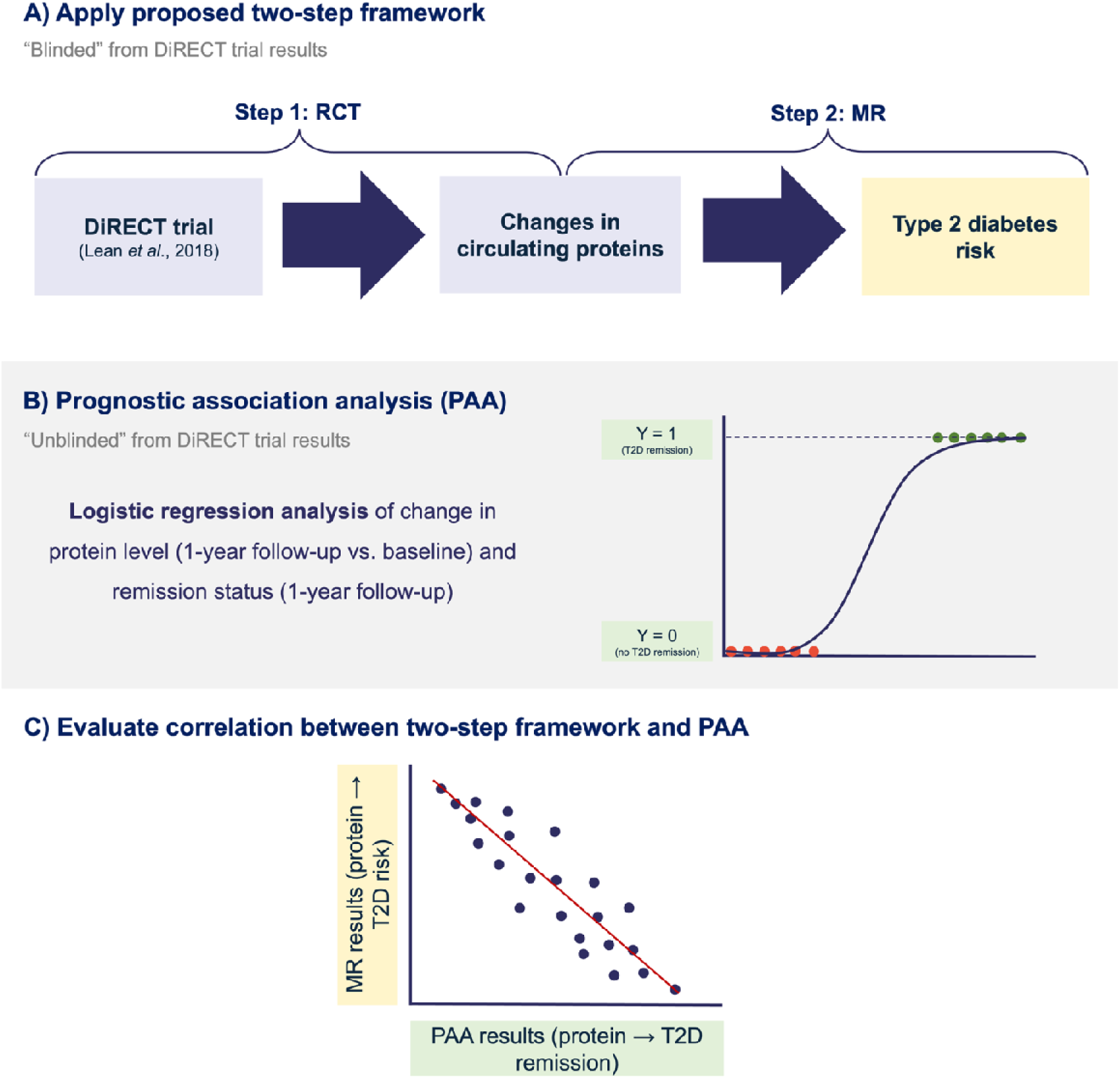
Schematic overview of the methodological framework. **(A)** Apply the two-step framework to DiRECT trial data (Lean et al., 2018), using proteins that changed in response to the dietary intervention relative to the control group (Step 1; analysis previously published by Goudswaard et al., 2023)(68), and estimating the effect of a 1-standard deviation (SD) change in protein levels on type 2 diabetes (T2D) risk using Mendelian randomisation (MR). **(B)** Using DiRECT trial data for a separate analysis: a logistic regression analysis to assess the association between changes in protein levels (follow-up vs baseline) and T2D remission at follow-up. **(C)** Evaluate correlation between effect estimates from the two-step framework (MR) and the trial-based prognostic association analysis (PAA).

### DiRECT study description

Data were obtained from the DiRECT trial (7), a cluster-RCT conducted in UK primary care that evaluated whether a structured weight management programme could induce remission of T2D. Participants were aged 20 to 65 years with a recent diagnosis of T2D (≤6 years) and a body mass index (BMI) between 27 and 45 kg/m^2^. General practitioner (GP) practices were randomised to either an intervention or control group. The intervention consisted of the Counterweight-Plus programme, which included an initial total diet replacement phase using a low-energy formula diet (∼850 kcal/day), followed by structured food reintroduction and ongoing weight maintenance support. Participants in the intervention group also discontinued glucose-lowering medications. The control group received best-practice care according to clinical guidelines. The intervention produced substantial weight loss and high rates of T2D remission at 12 months compared with standard care (46% vs 4% respectively) (7). Further details of the trial design and intervention have been described previously (7,67).

Clinical and biochemical measurements were collected at baseline and 1-year follow-up. Plasma proteins were quantified using the SomaScan v4 proteomic platform (SomaLogic), and 4,601 proteins passed quality control for downstream analyses (68). The primary outcome was T2D remission at 12 months, defined as HbA1c<48 mmol/mol in the absence of glucose-lowering medication (7).

DiRECT provides a useful example of the framework because the intervention produced large differences in both weight loss and T2D remission. The pathophysiological basis of T2D development and remission is now well characterised, although important questions remain regarding genetic susceptibility, including factors influencing the personal fat threshold and beta-cell susceptibility to fat-induced dysfunction (69,70). We can therefore compare protein-T2D risk effect estimates derived from the proposed two-step framework with protein-remission associations estimated directly within the trial using prognostic association analysis (PAA) (71). Although T2D remission and disease risk represent distinct clinical outcomes, remission cannot currently be examined with MR as there are no GWASs of T2D remission. For this validation analysis, we therefore assume that T2D risk and remission reflect overlapping biological pathways, such that factors increasing disease risk are directionally consistent with a lower probability of remission. This assumption is supported evidence that some factors associated with T2D development, including excess adiposity and beta cell dysfunction, also contribute to remission (7).

### (A) Applying the two-step framework

#### Step 1: identifying intervention-response intermediates

We used published results from Goudswaard et al., (2023) which quantified changes in circulating proteins following the DiRECT intervention (68). Here, differential protein levels were assessed using a group*timepoint interaction, identifying proteins whose changes over time differed between the intervention and control groups. 216 of 4,601 measured proteins showed changes (FDR-P<0.05). These proteins therefore represent intermediates linking the dietary intervention to downstream disease outcomes. The full list of these proteins is provided in Supplementary Table 1, which were taken forward to the next step of our analysis. Further details of the analysis pipeline have been described previously (68).

#### Step 2: Testing causal effects of intermediates on disease outcomes

To determine whether the 216 intervention-responsive proteins may causally influence T2D risk, we performed summary-level MR analyses using genetic variants strongly and independently associated with circulating protein levels as instrumental variables.

Genetic association data for circulating proteins were obtained from the proteomic GWAS conducted by Ferkingstad et al. (72) and T2D association data were obtained from the DIAGRAM consortium meta-analysis, which included 80,154 cases and 853,816 controls of European ancestry (73). We selected *cis-*protein quantitative trait loci (pQTLs) located within ±1 Mb of the protein-coding gene region. Variants associated with protein levels at genome-wide significance (P<5×10^-8^) were pruned for linkage disequilibrium (LD) (r^2^<0.001, 10 Mb window) using the 1000 Genomes Project Phase 3 reference panel (74) to identify independent instruments. Following instrument selection, suitable genetic instruments were available for 146 of the 216 proteins identified in Step 1 (Supplementary Table 2).

Exposure and outcome datasets were harmonised to ensure consistent effect allele orientation. Where an instrument was unavailable in the outcome dataset, proxy variants in high linkage disequilibrium (r^2^>0.8) were used where available. Summary-level MR analyses were conducted using the TwoSampleMR R package (75,76). Because only a single-SNP instruments were used for each protein, causal effects were estimated using the Wald ratio method, with standard errors calculated using the delta method (77). Multiple testing correction was applied using the Benjamini– Hochberg false discovery rate (FDR) (78).

We provide a non-exhaustive summary of commonly used GWAS resources for intermediate traits in Supplementary Table 3. Additional GWASs can be identified through public repositories (https://opengwas.io, https://www.ebi.ac.uk/gwas/ and http://www.metabolomix.com/category/resources/) and the wider literature.

#### Integrated score

One limitation of the two-step framework is that MR estimates represent the individual effect of a 1-SD change in an intermediate trait on disease risk. Consequently, the framework does not account for the magnitude of change, or collective effects, of protein level induced by the dietary intervention itself. To address this, we explored a novel summary metric which combines evidence from both steps of the framework – integrated score.

For each protein, the estimated effect of the dietary intervention on protein levels (Step 1) is multiplied by the corresponding MR estimate for the effect of that protein on disease risk (Step 2). These protein-specific effects were then combined using inverse-variance weighting to obtain an integrated score, with greater weight given to more precisely estimated effects.

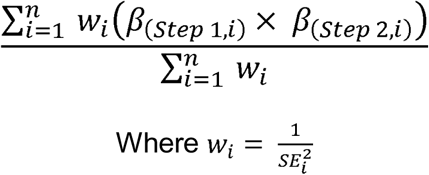

The integrated score is intended as an exploratory measure to summarise the overall direction and magnitude of the molecular effects of the intervention, rather than a formal estimate of the causal effect of the dietary intervention on disease risk.

### (B) Prognostic association analysis: Evaluating the framework using trial outcomes

To evaluate the validity of the two-step framework, we compared proteins identified as influencing T2D risk from MR with observed associations between protein changes and T2D remission within the DiRECT trial. This was achieved using PAA (71), in which changes in protein levels were related to remission status at 1-year follow-up.

Samples with excessive missing data and identified outliers using principal component analysis were excluded. Moreover, as established by Goudswaard et al., most proteins did not follow a normal distribution based on the Shapiro-Wilk test (W≥0.95) (68). Therefore, data was transformed prior to analysis using rank-based inverse transformation to standardise protein levels to have an approximate mean of 0 and a SD of 1.

Associations between changes in protein levels from baseline to 1-year follow-up, and T2D remission were assessed using mixed-effects logistic regression using the “lme4” R package. Analyses were conducted in the full sample, including both control and intervention participants. Models were adjusted for age, sex, list size, and study centre, with GP practice included as a random effect to account for clustering within practices. Variable definitions are provided in Supplementary Table 4. Multiple testing correction was applied using the Benjamini–Hochberg FDR (78).

### (C) Evaluating correlation between two-step framework and PAA estimates

Finally, we assessed the agreement between proteins identified through the two-step framework and the PAA analysis, using a correlation analysis.

## Results

### (A) Two-step framework

MR results are interpreted as the effect of a 1-SD increase in circulating protein levels on T2D risk. To aid interpretation, results are presented relative to the observed intervention effect. Thus, proteins that increased following the intervention are interpreted in relation to the effect of higher protein levels on T2D risk, whereas proteins that decreased following the intervention are interpreted in relation to the effect of lower protein levels on T2D risk.

Of the 146 proteins that were altered by the intervention (Step 1), 10 proteins had evidence of a causal effect on T2D risk (Step 2) following multiple testing correction (FDR-P<0.05). Of these, six showed intervention-induced changes consistent with a reduction in T2D risk. Concordance was defined as the direction of the intervention-induced change in protein level being consistent with the direction of its estimated effect on T2D risk. For instance, proteins that increased following the intervention were expected to be associated with lower T2D risk, whereas proteins that decreased were expected to be associated with higher T2D risk. Specifically, increased levels of NCAN, NEGR1, DUSP13, SHBG, and CDON were associated with lower T2D risk, and decreased levels of ADH1B were associated with lower T2D risk. On the other hand, decreased levels of BDH2, APOA4, and HTRA1 were associated with higher risk. Increased levels of CCDC126 were also associated with higher risk. Full MR results are shown in Supplementary Table 5 and **Figure 5**.

**Figure 5.**
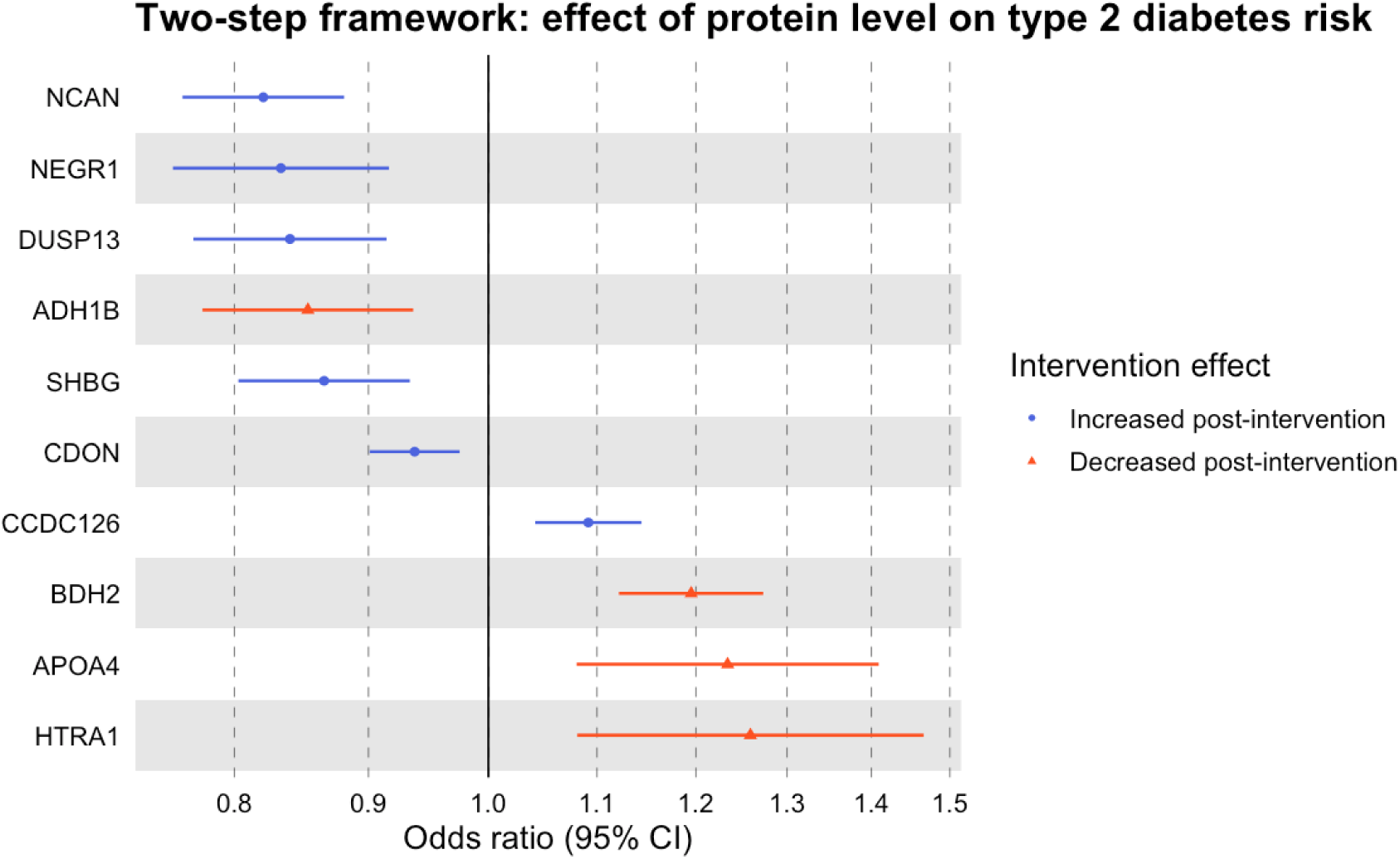
Forest plot of Mendelian randomisation (MR) estimates for circulating proteins and type 2 diabetes (T2D) risk. Odds ratios represent the estimated effect of a 1-standard deviation (SD) change in circulating protein levels on T2D risk. Proteins shown in red decreased following the dietary intervention at 1-year follow-up compared with the control group, whereas proteins shown in blue increased following the intervention. Because MR estimates are conventionally interpreted as the effect of increased exposure on disease risk, effect estimates for proteins that decreased following the intervention were directionally aligned (“flipped”) to reflect the direction of protein change observed in response to the intervention. Error bars represent 95% confidence intervals. Concordance was defined as the direction of the intervention-induced change in protein level being consistent with the direction of its estimated effect on T2D risk. For instance, proteins that increased following the intervention were expected to be associated with lower T2D risk, whereas proteins that decreased were expected to be associated with higher T2D risk.

#### Integrated score

The integrated score across the ten proteins was −0.0155 (SE = 0.007; 95% CI: −0.030 to −0.001; P = 0.04). This negative value indicates that the combined intervention-induced changes in circulating protein levels were overall associated with a predicted reduction in T2D risk. This direction of effect is consistent with the DiRECT trial findings (7), in which the dietary intervention substantially increased diabetes remission compared with standard care.

In a leave-one-out sensitivity analysis, the integrated score remained negative following the exclusion of each individual protein, although its magnitude varied. CCDC126 had the greatest influence on the overall estimate score (Supplementary Table 6, Supplementary Figure 1). Its exclusion strengthened the integrated score from −0.0155 to −0.0411 (95% CI: −0.059 to −0.024). Interestingly, CCDC126 was one of the two proteins for which the directions of the MR and PAA associations were discordant.

### (B) Prognostic association analysis

PAA identified 168 proteins associated with T2D remission at 1-year follow-up (FDR-P<0.05) (Supplementary Table 7; **Figure 6**).

**Figure 6.**
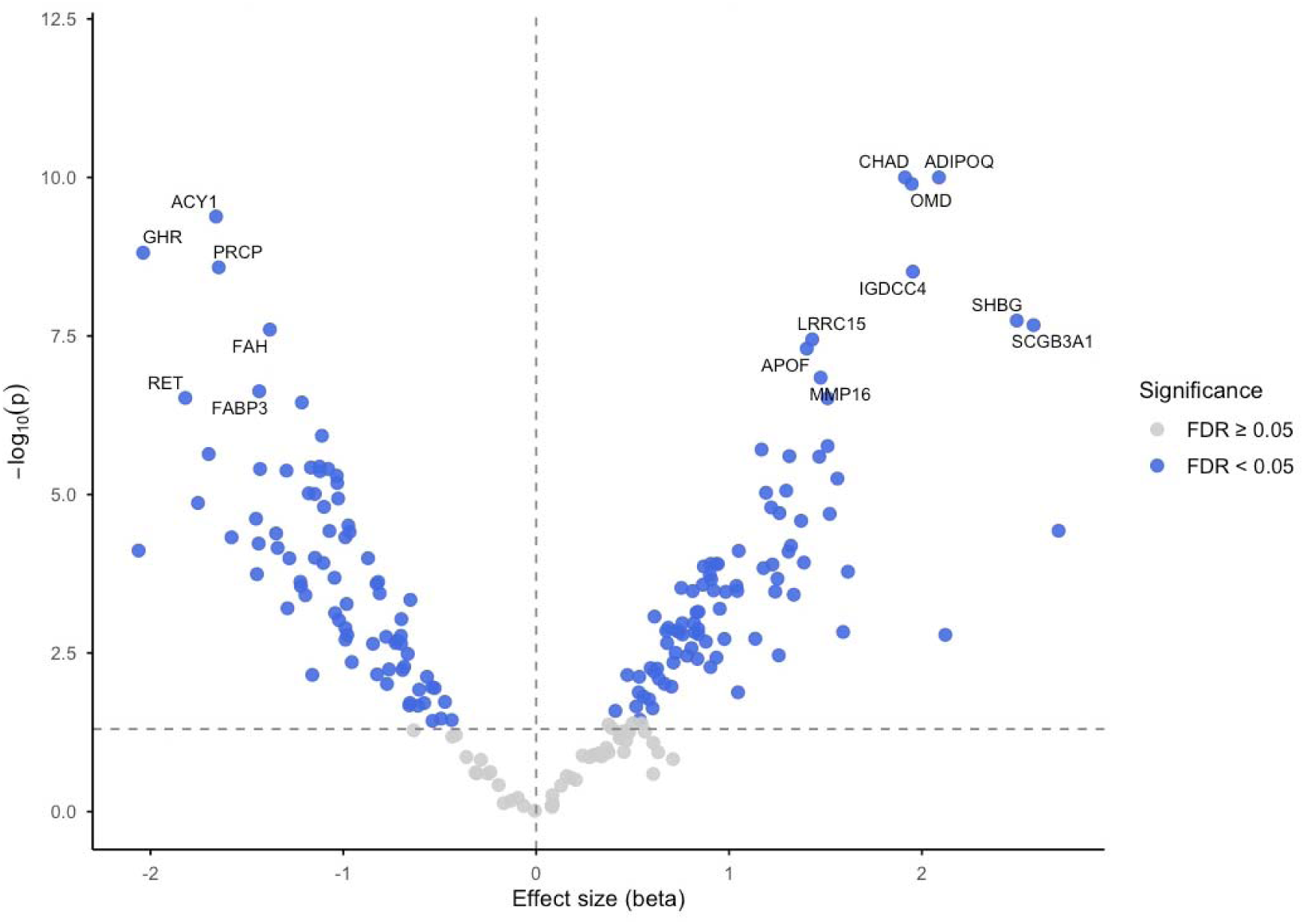
Volcano plot of prognostic association analysis (PAA) results from the DiRECT study. PAA was conducted using a mixed-effects logistic regression model to assess the association between changes in circulating protein levels (1-year follow-up versus baseline) and type 2 diabetes (T2D) remission status within the DiRECT trial (Lean et al., 2018). The x-axis represents the estimated effect size for the association between protein change and remission, and the y-axis represents the -log10 transformed P-value. Proteins shown in blue had FDR-P<0.05).

### (C) Evaluating correlation

Comparison of MR to trial-derived estimates is consistent with concordance between the two-step approach and RCT outcomes (r = −0.604, SE = 0.27; **Figure 7).** An inverse relationship is to be expected because MR estimates reflected effects on T2D risk, whereas PAA examined T2D remission. A comparison of protein-specific effect estimates from both approaches is shown in **Supplementary Figure 2.** Of the 10 proteins identified through the two-step framework, 8 showed concordant directional effects in both analyses. For example, proteins that increased following the intervention and were associated with lower T2D risk in the MR analysis also tended to be associated with a higher probability of remission in the trial-based analysis. Two proteins, ADH1B and CCDC126, showed discordant effects between the MR and PAA.

**Figure 7.**
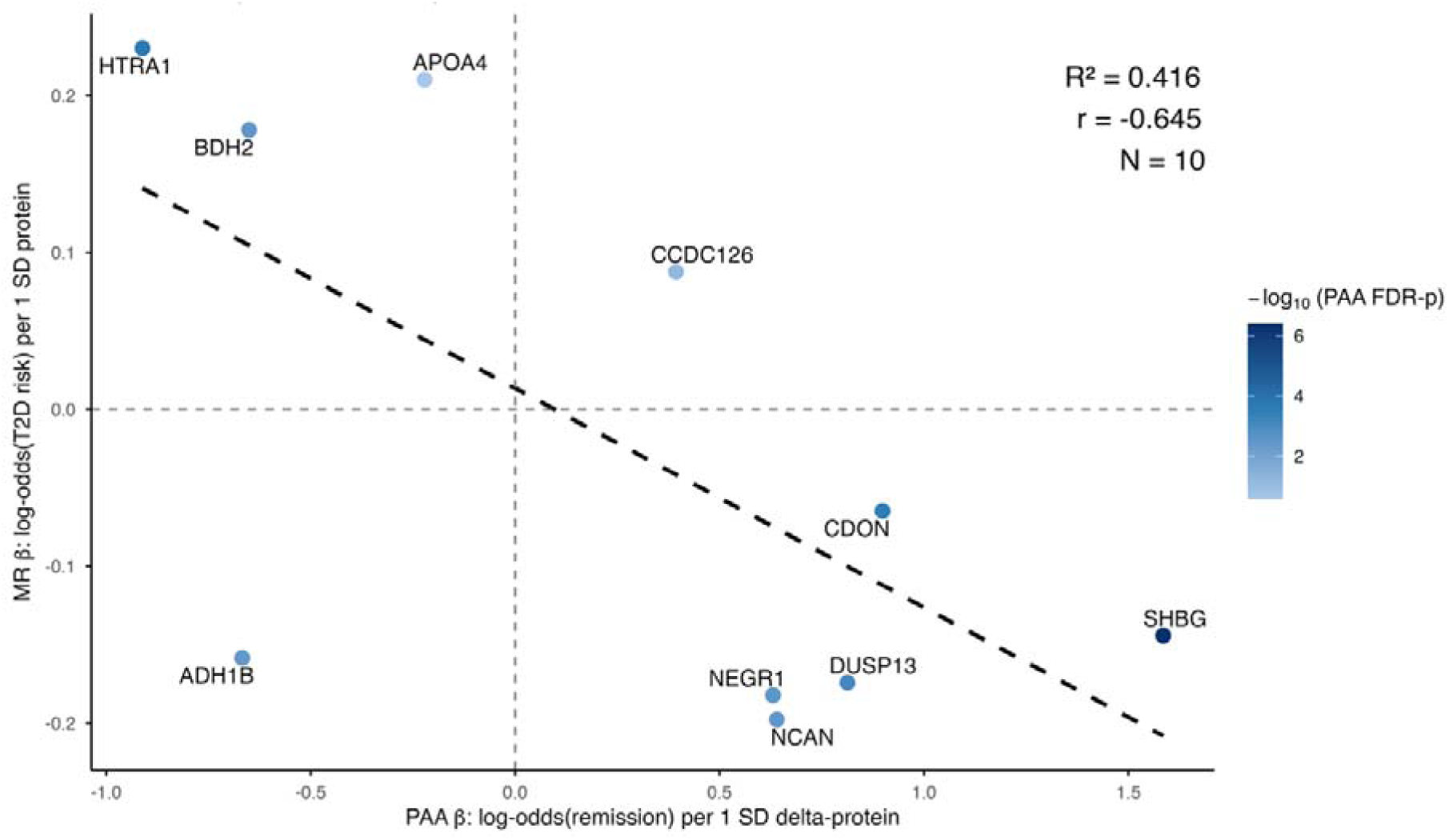
Scatter plot comparing effect estimates from the two-step framework and prognostic association analysis (PAA). Mendelian randomisation (MR) estimates represent the effect of a 1-SD change in circulating protein levels on type 2 diabetes (T2D) risk, whereas PAA estimates represent the association between a 1-SD increase in protein change from baseline to 1-year follow-up and T2D remission within the DiRECT trial (Lean et al., 2018). Each point represents an individual protein. Points are coloured according to the strength of the PAA results, with darker colours indicating stronger evidence of association.

## Discussion

Establishing the long-term causal effects of dietary interventions is challenging, as dietary RCTs are often limited in duration, while complex dietary exposures can be difficult to investigate using MR. Here we sought to bridge these complementary limitations by integrating short-term dietary intervention data with MR, in an approach we term the two-step framework. We find that the proposed two-step framework can identify intermediate molecular traits with potential causal effects on long-term outcomes, proving a means of linking diet and disease. Of the 10 proteins identified through the framework, 8 showed concordant associations when evaluated against remission outcomes within the DiRECT trial. The moderate inverse correlation observed between MR-derived estimates of T2D risk and trial-based estimates of T2D remission further supports the validity of the framework. Given that T2D risk and remission represent related but distinct outcomes, perfect agreement would not be expected.

Our findings are consistent with previous studies suggesting that short-term molecular responses to dietary exposures may capture signals relevant to longer-term disease outcomes. A similar conceptual approach (79) found that proteomic signatures associated with adherence to multiple dietary patterns, were subsequently associated with lower risk of T2D and other chronic illnesses. Similarly, metabolomic analysis from the DiRECT study (80) reported a strong inverse correlation between intervention-induced metabolite changes and metabolite associations with T2D risk. This supports the concept that intervention-responsive molecular traits may provide insight into disease-relevant biological processes.

Several proteins identified by the framework have previously been associated with T2D risk, including SHBG, BDH2, NCAN and HTRA1 (81–85), providing external support for the validity of the approach. However, the primary objective of this analysis was not to establish the role of individual proteins in T2D pathophysiology, but rather to assess whether the framework could identify and/or prioritise molecular intermediates that show evidence of relevance to disease outcomes.

Two proteins, ADH1B and CCDC126, did not show concordant findings between the MR and PAA. Interestingly, ADH1B expression in subcutaneous adipose tissue has previously been reported to have an inverse association with adiposity and insulin resistance, and also shown to increase following weight loss (86,87), whereas circulating ADH1B decreased following the intervention in DiRECT (68). This may reflect differences between tissue-specific expression and circulating protein levels, although the relationship between the two remains unclear. More broadly, discrepancies may reflect variation between the effects captured by MR and the trial-based analyses, limitations of the available genetic instruments, differences between T2D risk and remission as outcomes, or changes in protein levels that reflect the broader response to the intervention rather than causal intermediates. Distinguishing between these possibilities will require further investigation.

Taken together, these findings highlight the potential value of integrating RCT-derived molecular intermediates with MR. Dietary RCTs are often limited by short follow-up periods and therefore rely on intermediate outcomes, whereas MR alone is often unsuitable to provide robust evidence of causal relationships between dietary exposures and disease outcomes. By combining these complementary approaches, the two-step framework provides a practical approach for prioritising candidate endpoints for further investigation. Importantly, the framework is not intended to replace either RCTs or MR when used independently, but rather to strengthen causal inference by leveraging the complementary strengths of both approaches.

Several limitations of the proposed framework should be considered. First, estimates derived from RCTs and MR typically capture different causal contrasts. RCTs usually capture the effect of an intervention over a relatively short period of time, whereas MR estimates reflect the lifetime effect of an exposure. As a result, the magnitude and direction of effects may not be comparable. This is particularly relevant when interpreting intermediate traits, as short-term changes observed in an RCT may not reflect long-term biological adaptations (3,88). Thus, concordance between these approaches should be interpreted primarily in terms of direction and ranking of candidate endpoints rather than exact agreement in effect sizes. Where possible, findings should be triangulated against additional sources of evidence, including observational studies, mechanistic experiments, or replication in independent intervention studies.

Second, application of the framework assumes that MR estimates expressed per 1-SD increase in a trait can be interpreted in the opposite direction when the dietary intervention decreases that trait. This assumption may not always hold especially for non-linear relationships (89). In addition, intervention-induced molecular changes and MR estimates are not directly comparable because MR estimates are standardised to a 1-SD difference in the molecular trait, regardless of the size of the intervention effect. To partly address this limitation, we explored the use of an integrated score combining the magnitude of intervention protein changes with the corresponding MR estimates. This provides a useful summary of the overall direction of effects, but should not be interpreted as a causal estimate of the dietary intervention on disease risk, as it relies on assumptions regarding the comparability of RCT and MR estimates and may be sensitive to influential proteins. Further validation is required.

The framework also assumes that the intervention-disease relationship is captured, at least in part, through the individual intermediate trait under investigation. However, dietary interventions typically induce coordinated changes across tissues and many correlated molecular traits, making it difficult to distinguish causal intermediates from markers of broader biological processes (90,91). Consequently, proteins identified by the framework should be interpreted as prioritised candidate endpoints rather than definitive mediators of the intervention effect. Additional analyses, such as pathway enrichment, network analyses, multivariable MR, or triangulation with complementary evidence, may help distinguish independent mechanisms from correlated signals.

Third, the outcome evaluated in the trial (T2D remission) differed from the outcome used in the MR analysis (T2D risk). Although these are distinct outcomes, for this analysis we assume that T2D risk and remission reflect overlapping biological pathways, such that factors increasing disease risk are directionally consistent with a lower probability of remission. However, this assumption is imperfect given the substantial heterogeneity in T2D pathophysiology. For example, insulin resistance and insulin-signalling abnormalities vary considerably among individuals with T2D (92). Moreover, differences in study populations between the RCT and the GWAS used for MR may introduce heterogeneity. For example, the DiRECT trial has set eligibility criteria, whereas GWASs of T2D often capture a heterogeneous population with varying underlying pathophysiology, including differences in onset of diabetes, insulin resistance, beta-cell function, and adiposity. As a result, MR estimates may not be directly comparable to those observed in the trial population. When applying the framework, the user should carefully consider whether outcomes and population measured in intervention studies align sufficiently with those available in existing GWAS resources.

Finally, the framework is currently applicable to molecular intermediate traits, such as circulating proteins, metabolites, or gene expression, for which large-scale GWAS data and robust genetic instruments are available. Although the underlying principles could be extended to other intermediate phenotypes as appropriate genetic resources become available, application beyond molecular traits is currently limited. Furthermore, the DiRECT analysis should be viewed as a positive control example rather than conclusive validation of the framework. Although the concordance observed between the two-step framework and the trial-based analyses is encouraging, demonstrating robustness will require application across multiple interventions, molecular traits, and disease outcomes.

## Conclusion

We propose a two-step framework that integrates dietary intervention RCTs with MR through intermediate molecular traits. By linking short-term biological responses observed in dietary trials with long-term disease risk estimated using genetic instruments, this approach provides a complementary strategy for strengthening causal inference in nutrition research and prioritising candidate subclinical endpoints for further investigation. The framework is intended to complement, rather than replace, the primary objectives of dietary randomised controlled trials, and findings should be interpreted alongside the randomised trial outcomes and supported by additional evidence where possible. As RCT datasets and GWAS summary statistics become increasingly accessible, this approach may offer a practical and cost-effective framework for strengthening causal inference in nutrition research.

---

**Supplementary Figure 1.** Forest plot of leave-one-out sensitivity analysis of integrated score. The integrated score combines the intervention effect and Mendelian randomization (MR) effect for each protein to estimate the overall effect of the intervention-related protein changes on type 2 diabetes risk. For each protein, the intervention effect was multiplied by the MR effect, meaning that the direction of the score takes into account whether the protein increased or decreased following the intervention. A negative score therefore indicates that, overall, the intervention-related changes in protein levels are associated with lower type 2 diabetes risk, whereas a positive score indicates an association with higher type 2 diabetes risk. Protein-specific estimates were combined using inverse-variance weighting. The top row shows the integrated score using all proteins. The remaining rows show the integrated score after removing each protein in turn, to assess whether the overall result was strongly influenced by any single protein. Points show the integrated score and horizontal lines show 95% confidence intervals. The vertical grey line at zero represents no overall effect.

**Supplementary Figure 2.** Forest plot comparing effect estimates from Mendelian randomisation (MR) and prognostic association analysis (PAA). MR estimates (red) represent the effect of a 1-standard deviation (SD) change in circulating protein levels on type 2 diabetes (T2D) risk. PAA estimates (blue) represent the association between a 1-SD increase in protein change from baseline to 1-year follow-up and T2D remission within the DiRECT trial (Lean et al., 2018). As MR estimates reflect T2D risk whereas PAA estimates reflect T2D remission, effect estimates are expected to occur in opposite directions. Error bars represent 95% confidence intervals.

**Supplementary Table 1.** Circulating proteins altered following the DiRECT dietary intervention at 1-year follow-up (identified by Goudswaard et al.).

**Supplementary Table 2.** Proteins with available cis-pQTL genetic instruments included for MR analyses.

**Supplementary Table 3.** Summary of genome-wide association studies of molecular intermediate traits

**Supplementary Table 4**. Description of DiRECT study variables included in the prognostic association analysis.

**Supplementary Table 5.** Full MR results for circulating proteins and T2D risk.

**Supplementary Table 6.** Full leave-one-out analysis results.

**Supplementary Table 7.** Full results from mixed-effects logistic regression analysis of protein changes and T2D remission.

## Code and data availability

The code used to conduct the analyses reported in this study is publicly available on GitHub: https://github.com/bennydeslandes/twostepframework. The data underlying this study are not publicly available.

## Author contributions

B.D.: Conceptualization, Methodology, Formal analysis, Visualization, Writing - original draft, Writing - review & editing. L.J.C.: Conceptualization, Methodology, Formal analysis, Supervision, Writing - review & editing. L.J.G.: Conceptualization, Methodology, Formal analysis, Supervision, Writing - review & editing. M.R.S.: Writing - review & editing. M.A.L.: Formal analysis, Writing - review & editing. R.A.B.: Writing - review & editing. L.M.: Writing - review & editing. G.D.S.: Writing - review & editing. N.S.: Writing - review & editing. M.E.J.L.: Writing - review & editing. R.T.: Writing - review & editing. J.A.L.: Writing - review & editing. N.J.T.: Writing - review & editing. R.M.M.: Writing - review & editing. G.R.: Formal analysis, Writing - review & editing. M.J.G.: Writing - review & editing. J.Y.: Writing - review & editing. F.K.: Conceptualization, Supervision, Writing - review & editing. J.T.G.: Conceptualization, Supervision, Writing - review & editing. R.C.R.: Conceptualization, Methodology, Supervision, Writing - review & editing. E.E.V.: Conceptualization, Methodology, Supervision, Project administration, Writing - review & editing.

## Disclaimer

Where authors are identified as personnel of the International Agency for Research on Cancer/World Health Organization, the authors alone are responsible for the views expressed in this article and they do not necessarily represent the decisions, policy, or views of the International Agency for Research on Cancer/World Health Organization

## References

1. Merino J, Tobias DK. The unique challenges of studying the genetics of diet and nutrition. Nat Med. 2022 Feb;28(2):221–2. doi:10.1038/s41591-021-01626-w

2. Bowen DJ, Kreuter M, Spring B, Cofta-Woerpel L, Linnan L, Weiner D, et al. How we design feasibility studies. Am J Prev Med. 2009 May;36(5):452–7. doi:10.1016/j.amepre.2009.02.002 PubMed PMID: 19362699; PubMed Central PMCID: PMC2859314.

3. Smith GD, Ebrahim S. ‘Mendelian randomization’: can genetic epidemiology contribute to understanding environmental determinants of disease? Int J Epidemiol. 2003 Feb;32(1):1–22. doi:10.1093/ije/dyg070 PubMed PMID: 12689998.

4. Lawlor DA, Harbord RM, Sterne JAC, Timpson N, Davey Smith G. Mendelian randomization: Using genes as instruments for making causal inferences in epidemiology. Statistics in Medicine. 2008;27(8):1133–63. doi:10.1002/sim.3034

5. Genomic analysis of diet composition finds novel loci and associations with health and lifestyle | Molecular Psychiatry [Internet]. [cited 2025 Mar 3]. Available from: https://www.nature.com/articles/s41380-020-0697-5#additional-information

6. Smith GD, Hemani G, Ebrahim S. Gene-environment equivalence: The fundamental principle of Mendelian randomization. PLOS Medicine. 2026 Mar 13;23(3):e1005013. doi:10.1371/journal.pmed.1005013

7. Lean ME, Leslie WS, Barnes AC, Brosnahan N, Thom G, McCombie L, et al. Primary care-led weight management for remission of type 2 diabetes (DiRECT): an open-label, cluster-randomised trial. The Lancet. 2018 Feb 10;391(10120):541–51. doi:10.1016/S0140-6736(17)33102-1 PubMed PMID: 29221645.

8. David J. Torgerson and Carole J. Torgerson. Designing randomised trials in health, education and the social sciences: an introduction. England: Basingstoke, Hampshire; New York: Palgrave Macmillan 2008; 1960.

9. Prentice RL. Surrogate endpoints in clinical trials: Definition and operational criteria. Statistics in Medicine. 1989;8(4):431–40. doi:10.1002/sim.4780080407

10. DeMets DL, Psaty BM, Fleming TR. When Can Intermediate Outcomes Be Used as Surrogate Outcomes? JAMA. 2020 Mar 24;323(12):1184. doi:10.1001/jama.2020.1176

11. Sanderson E, Glymour MM, Holmes MV, Kang H, Morrison J, Munafò MR, et al. Mendelian randomization. Nat Rev Methods Primers. 2022 Feb 10;2(1):6. doi:10.1038/s43586-021-00092-5

12. Lawlor DA, Wade K, Borges MC, Palmer TM, Hartwig FP, Hemani G, et al. A Mendelian Randomization dictionary: Useful definitions and descriptions for undertaking, understanding and interpreting Mendelian Randomization studies [Internet]. Open Science Framework; 2019 [cited 2026 Jan 22]. Available from: https://osf.io/6yzs7_v1 doi:10.31219/osf.io/6yzs7

13. Davey Smith G, Hemani G. Mendelian randomization: genetic anchors for causal inference in epidemiological studies. Hum Mol Genet. 2014 Sep 15;23(R1):R89–98. doi:10.1093/hmg/ddu328

14. Wade KH, Yarmolinsky J, Giovannucci E, Lewis SJ, Millwood IY, Munafò MR, et al. Applying Mendelian randomization to appraise causality in relationships between nutrition and cancer. Cancer Causes Control. 2022 May 1;33(5):631–52. doi:10.1007/s10552-022-01562-1

15. Sanderson E, Rosoff D, Vitt N, Palmer T, Tilling K, Smith GD, et al. Heritable confounding in Mendelian randomization studies [Internet]. medRxiv; 2026 [cited 2026 Jun 22]. p. 2024.09.05.24312293. Available from: https://www.medrxiv.org/content/10.1101/2024.09.05.24312293v4 doi:10.1101/2024.09.05.24312293

16. Lippman SM, Klein EA, Goodman PJ, Lucia MS, Thompson IM, Ford LG, et al. Effect of Selenium and Vitamin E on Risk of Prostate Cancer and Other Cancers: The Selenium and Vitamin E Cancer Prevention Trial (SELECT). JAMA. 2009 Jan 7;301(1):39–51. doi:10.1001/jama.2008.864

17. Yarmolinsky J, Bonilla C, Haycock PC, Langdon RJQ, Lotta LA, Langenberg C, et al. Circulating Selenium and Prostate Cancer Risk: A Mendelian Randomization Analysis. J Natl Cancer Inst. 2018 Sep 1;110(9):1035–8. doi:10.1093/jnci/djy081

18. Mirmiran P, Bahadoran Z, Gaeini Z. Common Limitations and Challenges of Dietary Clinical Trials for Translation into Clinical Practices. Int J Endocrinol Metab. 2021 May 1;19(3):e108170. doi:10.5812/ijem.108170 PubMed PMID: 34567133; PubMed Central PMCID: PMC8453651.

19. Rothman Kj. INDUCTION AND LATENT PERIODS. Am J Epidemiol. 1981 Aug 1;114(2):253–9. doi:10.1093/oxfordjournals.aje.a113189

20. Schmitt J a. J, Bouzamondo H, Brighenti F, Kies AK, Macdonald I, Pfeiffer AFH, et al. The application of good clinical practice in nutrition research. Eur J Clin Nutr. 2012 Dec;66(12):1280–1. doi:10.1038/ejcn.2012.132

21. Weaver CM, Miller JW. Challenges in conducting clinical nutrition research. Nutr Rev. 2017 Jul 1;75(7):491–9. doi:10.1093/nutrit/nux026

22. Nichol AD, Bailey M, Cooper DJ. Challenging issues in randomised controlled trials. Injury. 2010 Jul;41:S20–3. doi:10.1016/j.injury.2010.03.033

23. Hróbjartsson A, Boutron I. Blinding in Randomized Clinical Trials: Imposed Impartiality. Clinical Pharmacology & Therapeutics. 2011;90(5):732–6. doi:10.1038/clpt.2011.207

24. Crichton GE, Howe PR, Buckley JD, Coates AM, Murphy KJ, Bryan J. Long-term dietary intervention trials: critical issues and challenges. Trials. 2012 Jul 20;13(1):111. doi:10.1186/1745-6215-13-111

25. Hewitt CE. Is there another way to take account of noncompliance in randomized controlled trials? Canadian Medical Association Journal. 2006 Aug 15;175(4):347–347. doi:10.1503/cmaj.051625

26. Byrd-Bredbenner C, Wu F, Spaccarotella K, Quick V, Martin-Biggers J, Zhang Y. Systematic review of control groups in nutrition education intervention research. Int J Behav Nutr Phys Act. 2017 Jul 11;14:91. doi:10.1186/s12966-017-0546-3 PubMed PMID: 28693581; PubMed Central PMCID: PMC5504837.

27. Robinson K, Allen F, Darby J, Fox C, Gordon AL, Horne JC, et al. Contamination in complex healthcare trials: the falls in care homes (FinCH) study experience. BMC Med Res Methodol. 2020 Feb 27;20(1):46. doi:10.1186/s12874-020-00925-z

28. Stender S, Gellert-Kristensen H, Smith GD. Reclaiming mendelian randomization from the deluge of papers and misleading findings. Lipids Health Dis. 2024 Sep 7;23(1):286. doi:10.1186/s12944-024-02284-w

29. Corbin LJ, Merino J, Fall T, Herder C. Raising the bar for publication of Mendelian randomisation studies in Diabetologia. Diabetologia. 2025 Oct 1;68(10):2088–91. doi:10.1007/s00125-025-06484-6

30. Hemani G, Stender S, Wolters FJ, Hofman A, Davey Smith G. The rapid growth in Mendelian randomization studies. Eur J Epidemiol. 2025 Oct 1;40(10):1165–71. doi:10.1007/s10654-025-01317-7

31. Hatcher C, McKinlay A, Bailward A, Dawes AC, Hughes DA, Pournaras DJ, et al. A systematic review of the applications of Mendelian randomization assessing the causal relevance of the gut microbiome in human health and disease [Internet]. medRxiv; 2025 [cited 2026 May 6]. p. 2025.06.03.25328787. Available from: https://www.medrxiv.org/content/10.1101/2025.06.03.25328787v1 doi:10.1101/2025.06.03.25328787

32. Burgess S, Thompson SG, CRP CHD Genetics Collaboration. Avoiding bias from weak instruments in Mendelian randomization studies. Int J Epidemiol. 2011 Jun;40(3):755– 64. doi:10.1093/ije/dyr036 PubMed PMID: 21414999.

33. Burgess S, Thompson SG. Bias in causal estimates from Mendelian randomization studies with weak instruments. Statistics in Medicine. 2011;30(11):1312–23. doi:10.1002/sim.4197

34. Glymour MM, Tchetgen Tchetgen EJ, Robins JM. Credible Mendelian Randomization Studies: Approaches for Evaluating the Instrumental Variable Assumptions. Am J Epidemiol. 2012 Feb 15;175(4):332–9. doi:10.1093/aje/kwr323

35. VanderWeele TJ, Tchetgen Tchetgen EJ, Cornelis M, Kraft P. Methodological Challenges in Mendelian Randomization. Epidemiology. 2014 May;25(3):427. doi:10.1097/EDE.0000000000000081

36. Burgess S, Butterworth A, Thompson SG. Mendelian Randomization Analysis With Multiple Genetic Variants Using Summarized Data. Genetic Epidemiology. 2013;37(7):658–65. doi:10.1002/gepi.21758

37. Brumpton B, Sanderson E, Heilbron K, Hartwig FP, Harrison S, Vie GÅ, et al. Avoiding dynastic, assortative mating, and population stratification biases in Mendelian randomization through within-family analyses. Nat Commun. 2020 Jul 14;11(1):3519. doi:10.1038/s41467-020-17117-4

38. Lawson DJ, Davies NM, Haworth S, Ashraf B, Howe L, Crawford A, et al. Is population structure in the genetic biobank era irrelevant, a challenge, or an opportunity? Hum Genet. 2020 Jan 1;139(1):23–41. doi:10.1007/s00439-019-02014-8

39. Sanderson E, Richardson TG, Hemani G, Davey Smith G. The use of negative control outcomes in Mendelian randomization to detect potential population stratification. Int J Epidemiol. 2021 Aug 1;50(4):1350–61. doi:10.1093/ije/dyaa288

40. Burgess S, Bowden J, Fall T, Ingelsson E, Thompson SG. Sensitivity Analyses for Robust Causal Inference from Mendelian Randomization Analyses with Multiple Genetic Variants. Epidemiology. 2017 Jan;28(1):30. doi:10.1097/EDE.0000000000000559

41. Bowden J, Davey Smith G, Haycock PC, Burgess S. Consistent Estimation in Mendelian Randomization with Some Invalid Instruments Using a Weighted Median Estimator. Genetic Epidemiology. 2016;40(4):304–14. doi:10.1002/gepi.21965

42. Hartwig FP, Davey Smith G, Bowden J. Robust inference in summary data Mendelian randomization via the zero modal pleiotropy assumption. Int J Epidemiol. 2017 Dec 1;46(6):1985–98. doi:10.1093/ije/dyx102

43. Zheng J, Baird D, Borges MC, Bowden J, Hemani G, Haycock P, et al. Recent Developments in Mendelian Randomization Studies. Curr Epidemiol Rep. 2017;4(4):330–45. doi:10.1007/s40471-017-0128-6 PubMed PMID: 29226067; PubMed Central PMCID: PMC5711966.

44. Morrison J, Knoblauch N, Marcus JH, Stephens M, He X. Mendelian randomization accounting for correlated and uncorrelated pleiotropic effects using genome-wide summary statistics. Nat Genet. 2020 Jul;52(7):740–7. doi:10.1038/s41588-020-0631-4

45. Mahmoud O, Dudbridge F, Davey Smith G, Munafo M, Tilling K. A robust method for collider bias correction in conditional genome-wide association studies. Nat Commun. 2022 Feb 2;13(1):619. doi:10.1038/s41467-022-28119-9

46. Paternoster L, Tilling K, Smith GD. Genetic epidemiology and Mendelian randomization for informing disease therapeutics: Conceptual and methodological challenges. PLOS Genetics. 2017 Oct 5;13(10):e1006944. doi:10.1371/journal.pgen.1006944

47. Gkatzionis A, Burgess S. Contextualizing selection bias in Mendelian randomization: how bad is it likely to be? Int J Epidemiol. 2019 Jun 1;48(3):691–701. doi:10.1093/ije/dyy202

48. Munafò MR, Tilling K, Taylor AE, Evans DM, Davey Smith G. Collider scope: when selection bias can substantially influence observed associations. Int J Epidemiol. 2018 Feb 1;47(1):226–35. doi:10.1093/ije/dyx206

49. Dudbridge F, Allen RJ, Sheehan NA, Schmidt AF, Lee JC, Jenkins RG, et al. Adjustment for index event bias in genome-wide association studies of subsequent events. Nat Commun. 2019 Apr 5;10(1):1561. doi:10.1038/s41467-019-09381-w

50. Watson JA, Leopold SJ, Simpson JA, Day NP, Dondorp AM, White NJ. Collider bias and the apparent protective effect of glucose-6-phosphate dehydrogenase deficiency on cerebral malaria. Lipsitch M, Ferguson NM, Murray E, editors. eLife. 2019 Jan 28;8:e43154. doi:10.7554/eLife.43154

51. Hartwig FP, Davies NM, Hemani G, Davey Smith G. Two-sample Mendelian randomization: avoiding the downsides of a powerful, widely applicable but potentially fallible technique. Int J Epidemiol. 2016 Dec 1;45(6):1717–26. doi:10.1093/ije/dyx028

52. Burgess S, Davies NM, Thompson SG. Bias due to participant overlap in two-sample Mendelian randomization. Genetic Epidemiology. 2016;40(7):597–608. doi:10.1002/gepi.21998

53. Fang S, Hemani G, Richardson TG, Gaunt TR, Davey Smith G. Evaluating and implementing block jackknife resampling Mendelian randomization to mitigate bias induced by overlapping samples. Hum Mol Genet. 2023 Jan 15;32(2):192–203. doi:10.1093/hmg/ddac186

54. Smith GD, Timpson N, Ebrahim S. Strengthening causal inference in cardiovascular epidemiology through Mendelian randomization. Annals of Medicine. 2008 Jan 1;40(7):524–41. doi:10.1080/07853890802010709

55. Staley JR, Burgess S. Semiparametric methods for estimation of a nonlinear exposure-outcome relationship using instrumental variables with application to Mendelian randomization. Genet Epidemiol. 2017 May;41(4):341–52. doi:10.1002/gepi.22041 PubMed PMID: 28317167; PubMed Central PMCID: PMC5400068.

56. Burgess S. Violation of the Constant Genetic Effect Assumption Can Result in Biased Estimates for Non-Linear Mendelian Randomization. Hum Hered. 2023 Aug 31;88(1):79–90. doi:10.1159/000531659

57. Tian H, Mason AM, Liu C, Burgess S. Relaxing parametric assumptions for non-linear Mendelian randomization using a doubly-ranked stratification method. PLOS Genetics. 2023 Jun 30;19(6):e1010823. doi:10.1371/journal.pgen.1010823

58. Burgess S, Davies NM, Thompson SG, Consortium on behalf of EI. Instrumental Variable Analysis with a Nonlinear Exposure–Outcome Relationship. Epidemiology. 2014 Nov;25(6):877. doi:10.1097/EDE.0000000000000161

59. Lousdal ML. An introduction to instrumental variable assumptions, validation and estimation. Emerg Themes Epidemiol. 2018 Jan 22;15(1):1. doi:10.1186/s12982-018-0069-7

60. Cortés J, González JA, Medina MN, Vogler M, Vilaró M, Elmore M, et al. Does evidence support the high expectations placed in precision medicine? A bibliographic review [Internet]. F1000Research; 2019 [cited 2026 Jan 12]. Available from: https://f1000research.com/articles/7-30 doi:10.12688/f1000research.13490.5

61. Winkelbeiner S, Leucht S, Kane JM, Homan P. Evaluation of Differences in Individual Treatment Response in Schizophrenia Spectrum Disorders: A Meta-analysis. JAMA Psychiatry. 2019 Oct 1;76(10):1063–73. doi:10.1001/jamapsychiatry.2019.1530

62. Senn S. Mastering variation: variance components and personalised medicine. Statistics in Medicine. 2016;35(7):966–77. doi:10.1002/sim.6739

63. Relton CL, Davey Smith G. Two-step epigenetic Mendelian randomization: a strategy for establishing the causal role of epigenetic processes in pathways to disease. Int J Epidemiol. 2012 Feb 1;41(1):161–76. doi:10.1093/ije/dyr233

64. Beynon RA, Richmond RC, Santos Ferreira DL, Ness AR, May M, Smith GD, et al. Investigating the effects of lycopene and green tea on the metabolome of men at risk of prostate cancer: The ProDiet randomised controlled trial. International Journal of Cancer. 2018;144(8):1918–28. doi:10.1002/ijc.31929

65. Bull CJ, Hazelwood E, Legge DN, Corbin LJ, Richardson TG, Lee M, et al. Impact of weight loss on cancer-related proteins in serum: results from a cluster randomised controlled trial of individuals with type 2 diabetes. eBioMedicine. 2024 Feb 1;100. doi:10.1016/j.ebiom.2024.104977 PubMed PMID: 38290287.

66. Pietzner M, Uluvar B, Kolnes KJ, Jeppesen PB, Frivold SV, Skattebo Ø, et al. Systemic proteome adaptions to 7-day complete caloric restriction in humans. Nat Metab. 2024 Apr;6(4):764–77. doi:10.1038/s42255-024-01008-9

67. Leslie WS, Ford I, Sattar N, Hollingsworth KG, Adamson A, Sniehotta FF, et al. The Diabetes Remission Clinical Trial (DiRECT): protocol for a cluster randomised trial. BMC Fam Pract. 2016 Feb 16;17(1):20. doi:10.1186/s12875-016-0406-2

68. Goudswaard LJ, Smith ML, Hughes DA, Taylor R, Lean M, Sattar N, et al. Using trials of caloric restriction and bariatric surgery to explore the effects of body mass index on the circulating proteome. Sci Rep. 2023 Nov 29;13(1):21077. doi:10.1038/s41598-023-47030-x

69. Taylor R, Al-Mrabeh A, Zhyzhneuskaya S, Peters C, Barnes AC, Aribisala BS, et al. Remission of Human Type 2 Diabetes Requires Decrease in Liver and Pancreas Fat Content but Is Dependent upon Capacity for β Cell Recovery. Cell Metabolism. 2018 Oct 2;28(4):547–556.e3. doi:10.1016/j.cmet.2018.07.003 PubMed PMID: 30078554.

70. Al-Mrabeh A, Zhyzhneuskaya SV, Peters C, Barnes AC, Melhem S, Jesuthasan A, et al. Hepatic Lipoprotein Export and Remission of Human Type 2 Diabetes after Weight Loss. Cell Metabolism. 2020 Feb 4;31(2):233–249.e4. doi:10.1016/j.cmet.2019.11.018 PubMed PMID: 31866441.

71. Simon RM, Paik S, Hayes DF. Use of Archived Specimens in Evaluation of Prognostic and Predictive Biomarkers. JNCI Journal of the National Cancer Institute. 2009 Nov 4;101(21):1446–52. doi:10.1093/jnci/djp335

72. Ferkingstad E, Sulem P, Atlason BA, Sveinbjornsson G, Magnusson MI, Styrmisdottir EL, et al. Large-scale integration of the plasma proteome with genetics and disease. Nat Genet. 2021 Dec;53(12):1712–21. doi:10.1038/s41588-021-00978-w

73. Mahajan A, Spracklen CN, Zhang W, Ng MCY, Petty LE, Kitajima H, et al. Multi-ancestry genetic study of type 2 diabetes highlights the power of diverse populations for discovery and translation. Nat Genet. 2022 May;54(5):560–72. doi:10.1038/s41588-022-01058-3

74. Auton A, Abecasis GR, Altshuler DM, Durbin RM, Abecasis GR, Bentley DR, et al. A global reference for human genetic variation. Nature. 2015 Oct;526(7571):68–74. doi:10.1038/nature15393

75. Hemani G, Zheng J, Elsworth B, Wade KH, Haberland V, Baird D, et al. The MR-Base platform supports systematic causal inference across the human phenome. eLife. 2018 May 30;7:e34408. doi:10.7554/eLife.34408

76. Hemani G, Tilling K, Davey Smith G. Orienting the causal relationship between imprecisely measured traits using GWAS summary data. Li J, editor. PLoS Genet. 2017 Nov 17;13(11):e1007081. doi:10.1371/journal.pgen.1007081

77. Hemani G, Bowden J, Davey Smith G. Evaluating the potential role of pleiotropy in Mendelian randomization studies. Hum Mol Genet. 2018 Aug 1;27(R2):R195–208. doi:10.1093/hmg/ddy163

78. Benjamini Y, Hochberg Y. Controlling the False Discovery Rate: A Practical and Powerful Approach to Multiple Testing. Journal of the Royal Statistical Society Series B (Methodological). 1995;57(1):289–300.

79. Zhu K, Li R, Yao P, Yu H, Pan A, Manson JE, et al. Proteomic signatures of healthy dietary patterns are associated with lower risks of major chronic diseases and mortality. Nat Food. 2025 Jan;6(1):47–57. doi:10.1038/s43016-024-01059-x

80. Corbin LJ, Hughes DA, Bull CJ, Vincent EE, Smith ML, McConnachie A, et al. The metabolomic signature of weight loss and remission in the Diabetes Remission Clinical Trial (DiRECT). Diabetologia. 2024 Jan 1;67(1):74–87. doi:10.1007/s00125-023-06019-x

81. Gupte TP, Azizi Z, Kho PF, Zhou J, Nzenkue K, Chen ML, et al. Plasma proteomic signatures for type 2 diabetes and related traits in the UK Biobank cohort. Diabetes Research and Clinical Practice. 2025 Jun 1;224. doi:10.1016/j.diabres.2025.112194 PubMed PMID: 40274105.

82. Elhadad MA, Jonasson C, Huth C, Wilson R, Gieger C, Matias P, et al. Deciphering the Plasma Proteome of Type 2 Diabetes. Diabetes. 2020 Dec;69(12):2766–78. doi:10.2337/db20-0296 PubMed PMID: 32928870; PubMed Central PMCID: PMC7679779.

83. Rooney MR, Chen J, Echouffo-Tcheugui JB, Walker KA, Schlosser P, Surapaneni A, et al. Proteomic Predictors of Incident Diabetes: Results From the Atherosclerosis Risk in Communities (ARIC) Study. Diabetes Care. 2023 Apr;46(4):733–41. doi:10.2337/dc22-1830 PubMed PMID: 36706097; PubMed Central PMCID: PMC10090896.

84. Yuan S, Xu F, Li X, Chen J, Zheng J, Mantzoros CS, et al. Plasma proteins and onset of type 2 diabetes and diabetic complications: Proteome-wide Mendelian randomization and colocalization analyses. Cell Rep Med. 2023 Aug 30;4(9):101174. doi:10.1016/j.xcrm.2023.101174 PubMed PMID: 37652020; PubMed Central PMCID: PMC10518626.

85. Ferrannini G, Manca ML, Magnoni M, Andreotti F, Andreini D, Latini R, et al. Coronary Artery Disease and Type 2 Diabetes: A Proteomic Study. Diabetes Care. 2020 Jan 27;43(4):843–51. doi:10.2337/dc19-1902

86. He H, Sun D, Zeng Y, Wang R, Zhu W, Cao S, et al. A Systems Genetics Approach Identified GPD1L and its Molecular Mechanism for Obesity in Human Adipose Tissue. Sci Rep. 2017 May 11;7:1799. doi:10.1038/s41598-017-01517-6 PubMed PMID: 28496128; PubMed Central PMCID: PMC5431993.

87. Morales LD, Cromack DT, Tripathy D, Fourcaudot M, Kumar S, Curran JE, et al. Further evidence supporting a potential role for ADH1B in obesity. Sci Rep. 2021 Jan 21;11(1):1932. doi:10.1038/s41598-020-80563-z

88. Richmond RC, Davey Smith G. Mendelian Randomization: Concepts and Scope. Cold Spring Harb Perspect Med. 2022 Jan;12(1):a040501. doi:10.1101/cshperspect.a040501 PubMed PMID: 34426474; PubMed Central PMCID: PMC8725623.

89. Kohlmeier M, Baah E. When Mendelian randomisation fails. BMJNPH. 2021 Jun;4(1):1– 3. doi:10.1136/bmjnph-2021-000265

90. Bogaards FA, Gehrmann T, Beekman M, Lakenberg N, Suchiman HED, de Groot CPGM, et al. Secondary integrated analysis of multi-tissue transcriptomic responses to a combined lifestyle intervention in older adults from the GOTO nonrandomized trial. Nat Commun. 2024 Aug 15;15(1):7013. doi:10.1038/s41467-024-50693-3

91. Valsesia A, Chakrabarti A, Hager J, Langin D, Saris WHM, Astrup A, et al. Integrative phenotyping of glycemic responders upon clinical weight loss using multi-omics. Sci Rep. 2020 Jun 8;10(1):9236. doi:10.1038/s41598-020-65936-8

92. Larsen JK, Stocks B, Henderson J, Andersson D, Bäckdahl J, Eriksson-Hogling D, et al. Personalized Molecular Signatures of Insulin Resistance and Type 2 Diabetes [Internet]. Biochemistry; 2024 [cited 2026 Aug 25]. Available from: http://biorxiv.org/lookup/doi/10.1101/2024.02.06.578994 doi:10.1101/2024.02.06.578994

