## Supplementary figures and images for "Connecting diet and disease: Using Mendelian randomisation to bridge the gap"

### Supplementary Figure 1

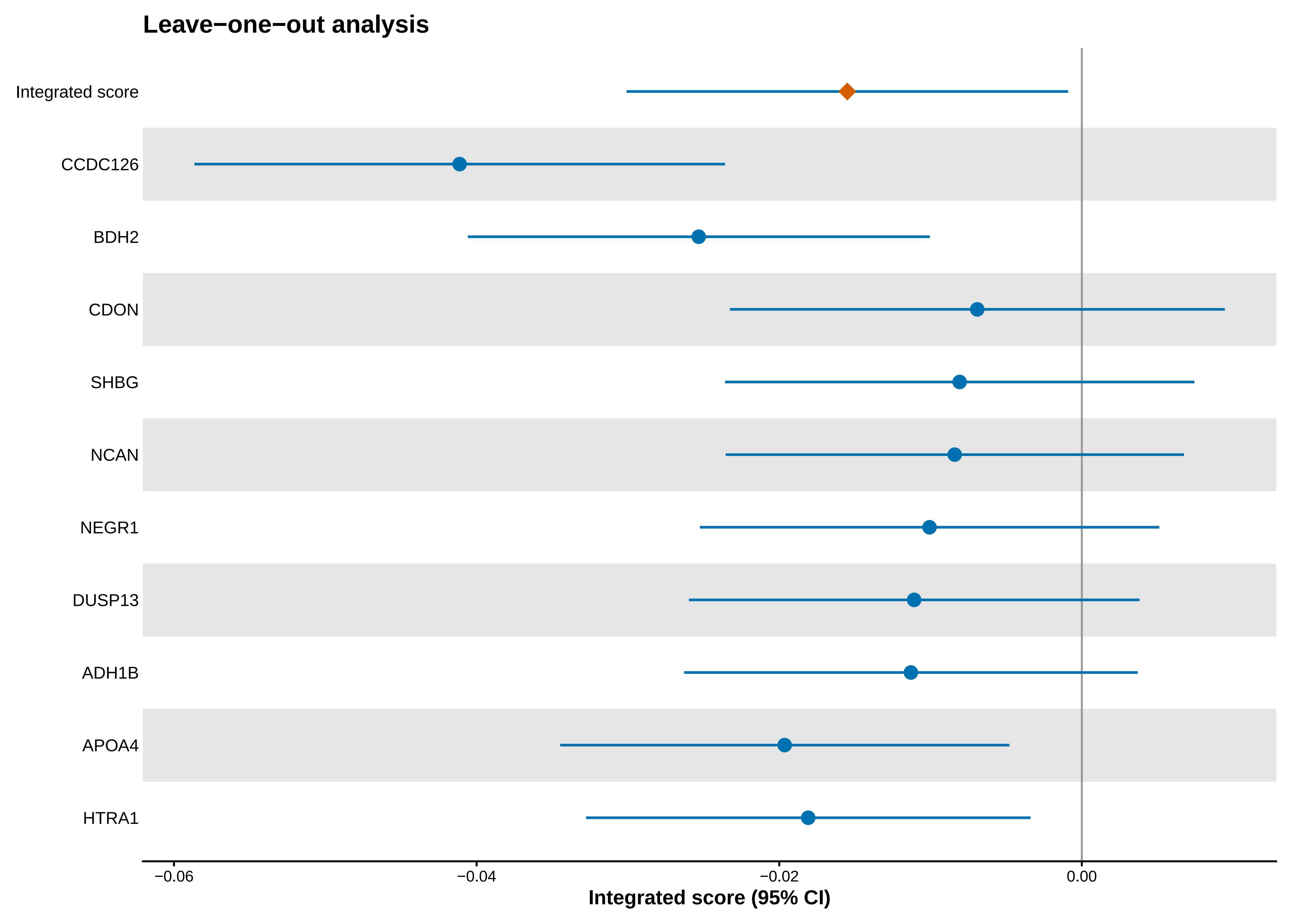

### Supplementary Figure 2

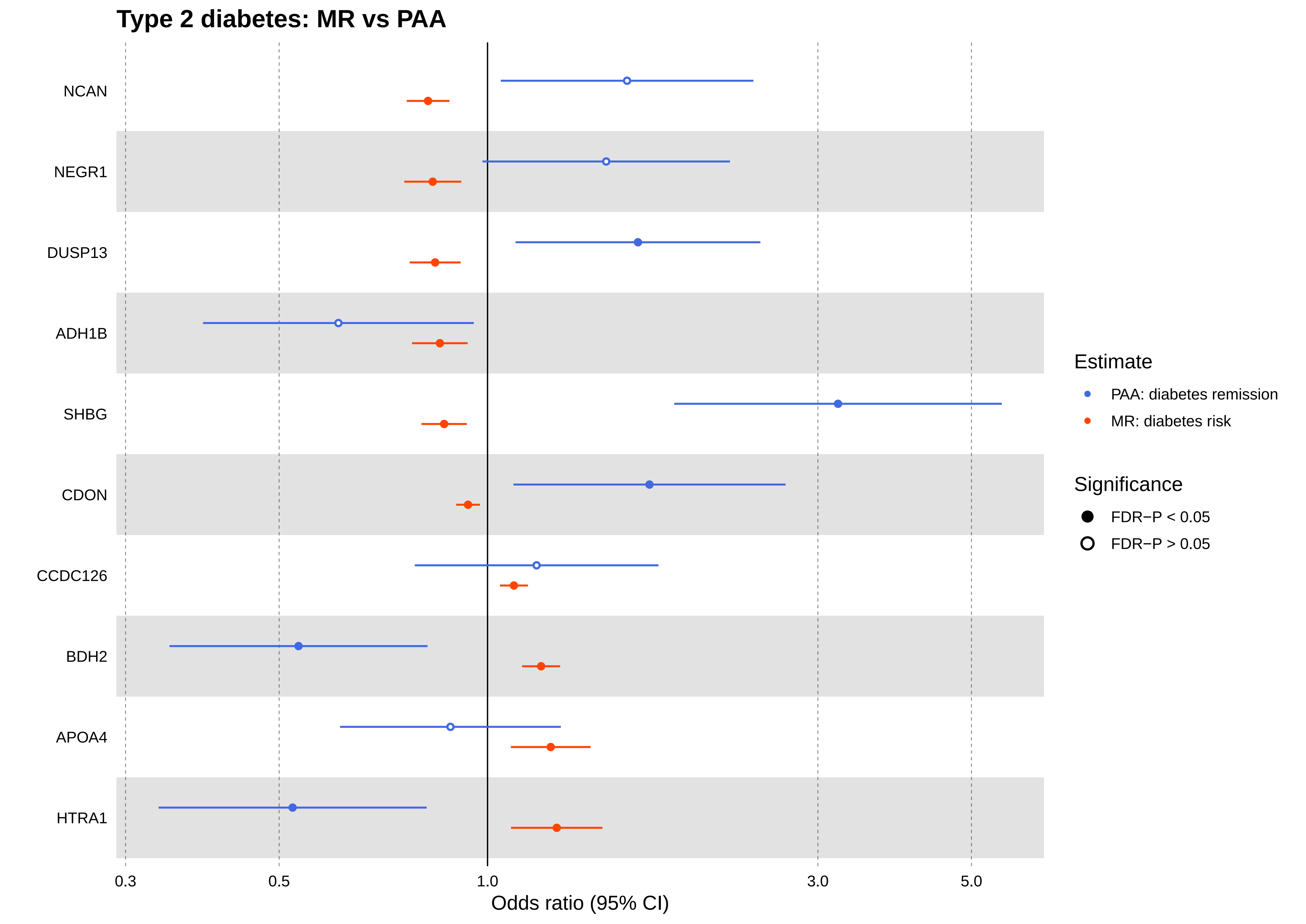
